# Sacubitril-valsartan Versus Enalapril or Losartan at Guideline-Recommended Maximum Dosages in Heart Failure with Reduced Ejection Fraction: Real-World Results from the BEAT-HF Cohort

**DOI:** 10.64898/2025.12.07.25341800

**Authors:** Renato C. Barros, Vinícius C. Fiusa, Gabrielle N. Tim, Victoria T. V. Goulart, Caio Henrique M. Nascimento, Gustavo T. de Moura, Adriana J. B. A. Guimarães, Sergio Henrique Rodolpho Ramalho, Armindo Jreige, Filipe A. Moura, Luiz Sérgio F. de Carvalho

## Abstract

**Background:** Sacubitril-valsartan reduced mortality and hospitalizations in patients with heart failure with reduced ejection fraction (HFrEF) compared to enalapril in pivotal clinical trials. However, these studies used suboptimal dosages of enalapril, potentially limiting their findings.

**Objective:** To compare the outcomes of HFrEF patients treated with sacubitril-valsartan versus either enalapril or losartan at guideline-recommended maximum dosages.

**Methods:** We analyzed data from BEAT-HF, an observational cohort of 2,314 HF patients who received care between Aug/2010 and Jan/2024 at six public hospitals in the Brasília, Brazil. The primary analysis included 254 HFrEF patients receiving the guideline-recommended maximum dosages of the investigated drugs. A sensitivity analysis included 452 patients receiving any sub-maximum dosages. The primary outcome was combined hospitalization or death. Baseline differences were adjusted using propensity score matching.

**Results:** Among 254 HFrEF patients (mean age 63.6±12.3 years, 61.4% male), 115 received sacubitril-valsartan 97/103mg twice daily, 33 enalapril 20mg twice daily, and 106 losartan 100mg daily. Over the median follow-up of 23.9 months [IQR 11.9 – 41.1], there were 70 primary endpoint events (56 first hospitalizations, 32 deaths). Compared to equivalent dosages of enalapril or losartan, sacubitril-valsartan was not associated with significantly reductions in combined hospitalization or death (OR 0.650, 95%CI 0.354 – 1.195, p=0.165), but it was strongly associated with functional status improvement (OR 3.902, 95%CI 1.745 – 8.726, p=0.001).

**Conclusion:** This real-world study wasn’t able to support significant reductions in hospitalization or mortality for sacubitril-valsartan compared with enalapril or losartan at guideline-recommended maximum dosages, suggesting that its benefits may be more modest than previously reported.

## INTRODUCTION

In the past decade, angiotensin receptor-neprilysin inhibitors (ARNIs) have emerged as a cornerstone treatment of heart failure with reduced ejection fraction (HFrEF). Sacubitril’s formulation with valsartan promote vasodilation and natriuresis while counteracting the cardiotoxic effects of angiotensin II [1].

Current HF guidelines, recommend ARNIs over angiotensin-converting enzyme (ACE) inhibitors or angiotensin receptor blockers (ARBs) for patients with HFrEF [2,3]. This recommendation was initially supported by the PARADIGM-HF trial, which demonstrated a significant reduction in mortality and hospitalizations with sacubitril-valsartan compared to enalapril [4]. Subsequently, a pooled analysis of data from the PIONEER-HF and PARAGLIDE-HF trials provided further evidence supporting the use of ARNIs in acute settings, addressing a key gap left by the run-in period of the PARADIGM-HF trial [5–7]. However, a common criticism of these studies lies in the use of a suboptimal dosage of enalapril (10mg twice daily, rather than the titratable maximum of 20mg twice daily, generally recommended by HF guidelines), compared to a full dosage of sacubitril-valsartan (97/103mg twice daily).

To address this controversy and provide a more equitable comparison, the present study aims to compare the clinical outcomes of patients with HFrEF treated with the guideline-recommended maximum dosages of sacubitril-valsartan (97/103mg twice daily) versus either enalapril (20mg twice daily) or losartan (100mg daily). This effort seeks to clarify the true benefits of ARNIs in a real-world clinical setting.

## METHODS

### Study design, population, and ethics

This study analyzed data from BEAT-HF (Brasilia Evaluation and Tracking of Heart Failure), an ongoing observational cohort of 2,314 patients with heart failure who received care between August 2010 and September 2024 at six public hospitals in the Brasilia metropolitan area (Hospital Regional do Guará, Hospital de Base do Distrito Federal, Hospital Regional de Sobradinho, Hospital Regional do Gama, Hospital Regional de Taguatinga, and Hospital Regional Leste). Based on the last national demographic census and on previous epidemiological HF studies in Brazil, BEAT-HF encompassed approximately 8.4% of all individuals with heart failure in the Brasília metropolitan area [8,9].

BEAT-HF enrolled patients 18 years and older who met the Framingham criteria for HF [10]. From January 2021 until January 2024, a total of 931 patients with HF were enrolled prospectively through structured clinical interview during outpatient follow-up, hospital visits for cardiac imaging studies, and hospitalizations. From October 2023 to September 2024, another 1,383 patients with HF registered at the local referral center (Hospital Regional do Guará) were enrolled retrospectively through digital medical records review. Patients had their medical history assessed since the diagnosis of HF, which was considered the participants’ baseline, with follow-up data from August 2010 to September 2024. During data extraction and review, we had access to information that could identify individual participants.

From the 2,314 patients in BEAT-HF, the current study only analyzed those with reduced ejection fractions (≤40%) (n=903). Moreover, individuals were excluded due to the following reasons: unavailable or incomplete prescription data (n=180); use of another type of angiotensin-converting enzyme (ACE) inhibitor or angiotensin receptor blocker (ARB) (n=17); and use of sub-maximal dosages of the investigated drugs (n=452). The final cohort for the primary analysis encompassed 254 eligible patients, as outlined in the STROBE flowchart (**Figure 1**). A sensitivity analysis, including the 452 patients on any sub-maximum dosages of the investigated drugs, was also performed.

**Figure 1.**
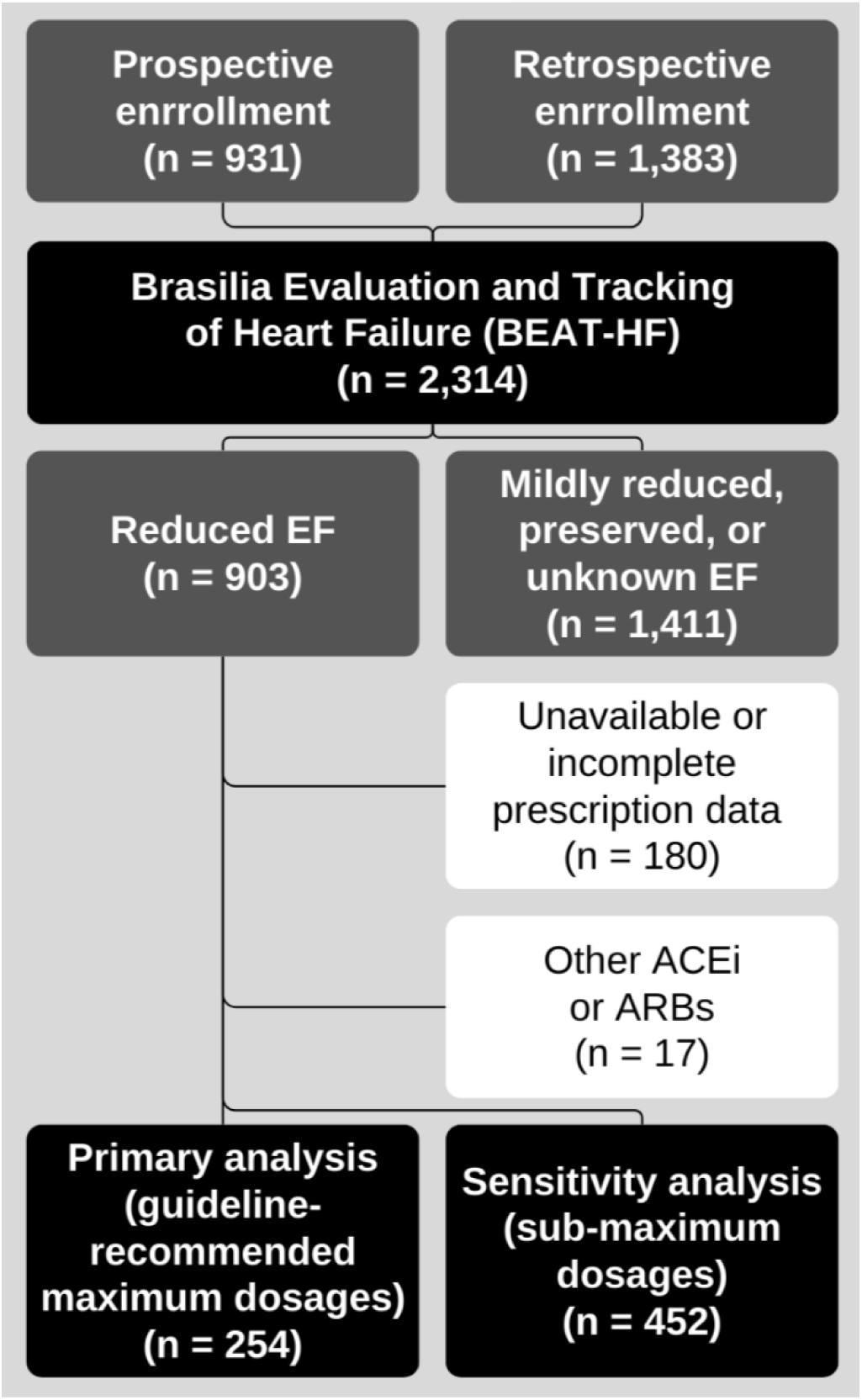
STROBE flowchart. EF: ejection fraction. ACEi: Angiotensin-Converting Enzyme Inhibitor. ARB: Angiotensin II Receptor Blocker.

The study methodology was approved by the Institutional Review Boards (IRBs) of Instituto de Gestão Estratégica de Saúde do Distrito Federal (IGESDF; approval number 35543020.1.0000.8153), and Fundação de Ensino e Pesquisa em Ciências de Saúde (FEPECS; approval number 72838823.5.0000.5553). Informed consent was obtained from all participants by physical or digital means, except for retrospectively enrolled participants who either died during follow-up or were deemed unreachable after two telephone contact attempts on different days, for whom a waiver was granted by the IRBs.

### Data collection and definitions

The REDCap platform was used for data capture. The following data categories were collected: demographics, comorbidities, pharmacotherapy, laboratory parameters, and clinical outcomes. The primary outcome was combined all-cause hospitalization or death. Secondary outcomes included three-point major adverse cardiovascular events (3p-MACE; defined as combined non-fatal myocardial infarction, non-fatal stroke, or cardiovascular death), and New York Heart Association (NYHA) class progression (no change, improvement, or worsening).

### Statistical analysis

Statistical analyses were conducted on RStudio version 4.4.2 for Windows. Categorical variables were represented as counts and percentages. Continuity-corrected Pearson’s chi-squared test was used to compare categorical variables with ≥10 occurrences, and Fisher’s exact test for those with <10 occurrences. Continuous variables with <50 cases had their normality evaluated by the Shapiro-Wilk test, and those with ≥50 cases by graphical inspection (histogram, Q-Q plot, and box plot). Normal continuous variables were represented as mean and standard deviation and compared using variance-adjusted independent Student T test. Non-normal continuous variables were represented as median and interquartile range (IQR) and compared using the Mann-Whitney test.

To account for baseline differences between treatment groups, inherent to observational studies, we performed propensity score matching (PSM). To evaluate the impact of sacubitril-valsartan on specific outcomes, we estimated the average treatment effect on the treated (ATET). We performed Multivariate imputation to improve statistical compatibility with these strategies (by minimizing the number of cases excluded due to missing covariates).

Multivariate imputation was conducted with the mice package. Continuous and binary variables with up to 20% missing data were imputed, except for: sex, age, outcomes, and the inclusion criteria (left ventricular ejection fraction, sacubitril-valsartan use, enalapril use, and losartan use). Logistic regression was used for the imputation of binary variables, and predictive mean matching for continuous variables. Categorical variables with more than 2 categories were not imputed due to the extreme distribution in most of them (ie, very few asians and indigenous in the race variable). This process reduced the already low percentage of missing cells in imputed variables from 3.53 to 0.42%. Rubin’s rules were applied to pool p-values across imputations in subsequent analyses.

Given the large number of variables in our dataset, LASSO regression (least absolute shrinkage and selection operator) was employed to guide covariate selection for PSM. The MatchIt package was used to perform PSM through the genetic method, chosen by its robustness and efficiency. ATET was estimated by weighted logistic regression and was restricted to events with more than 10 occurrences.

## RESULTS

From August 2010 and September 2024, 254 patients with HFrEF received care across six public hospitals in the Brasilia metropolitan area. A total of 115 patients (45.3%) were prescribed sacubitril-valsartan 97/103mg twice daily, 33 (13.0%) enalapril 20mg twice daily, and 106 (41.7%) losartan 100mg daily. The median follow-up of this cohort was 23.9 months [IQR 11.9 – 41.1].

### Population characteristics

The population was 61.4% male, with a mean age of 63.6 ± 12.3 years. If stratified by treatment, no significant differences were observed for sex, age, body composition, or race (**Table 1**). Comorbidities were also largely similar between groups. Individuals on sacubitril-valsartan had a lower LVEF (Mdn 29.0% [IQR 23.0 – 35.0] vs 33.0% [IQR 27.0 – 37.0], p<0.001).

**Table 1.**
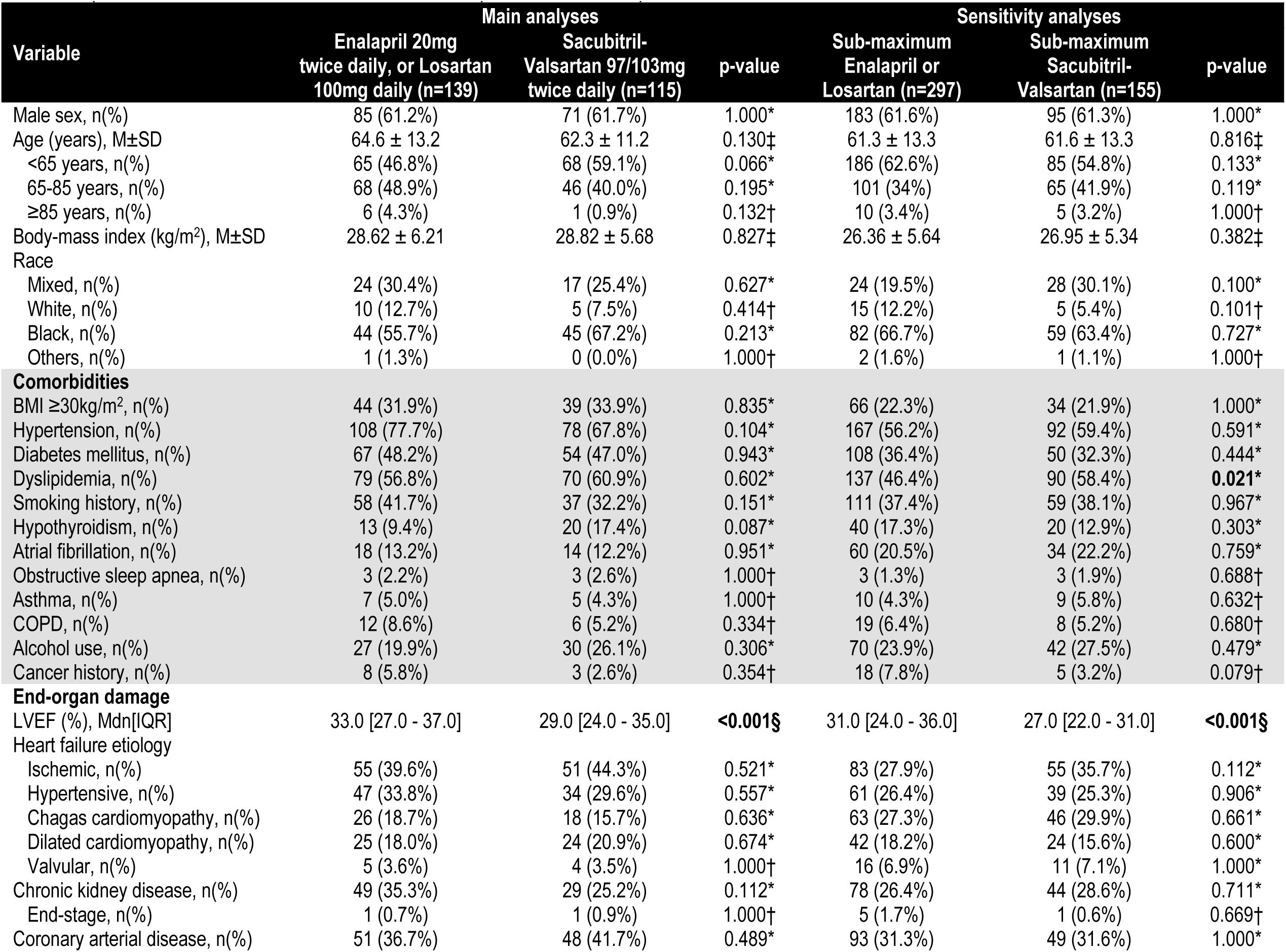
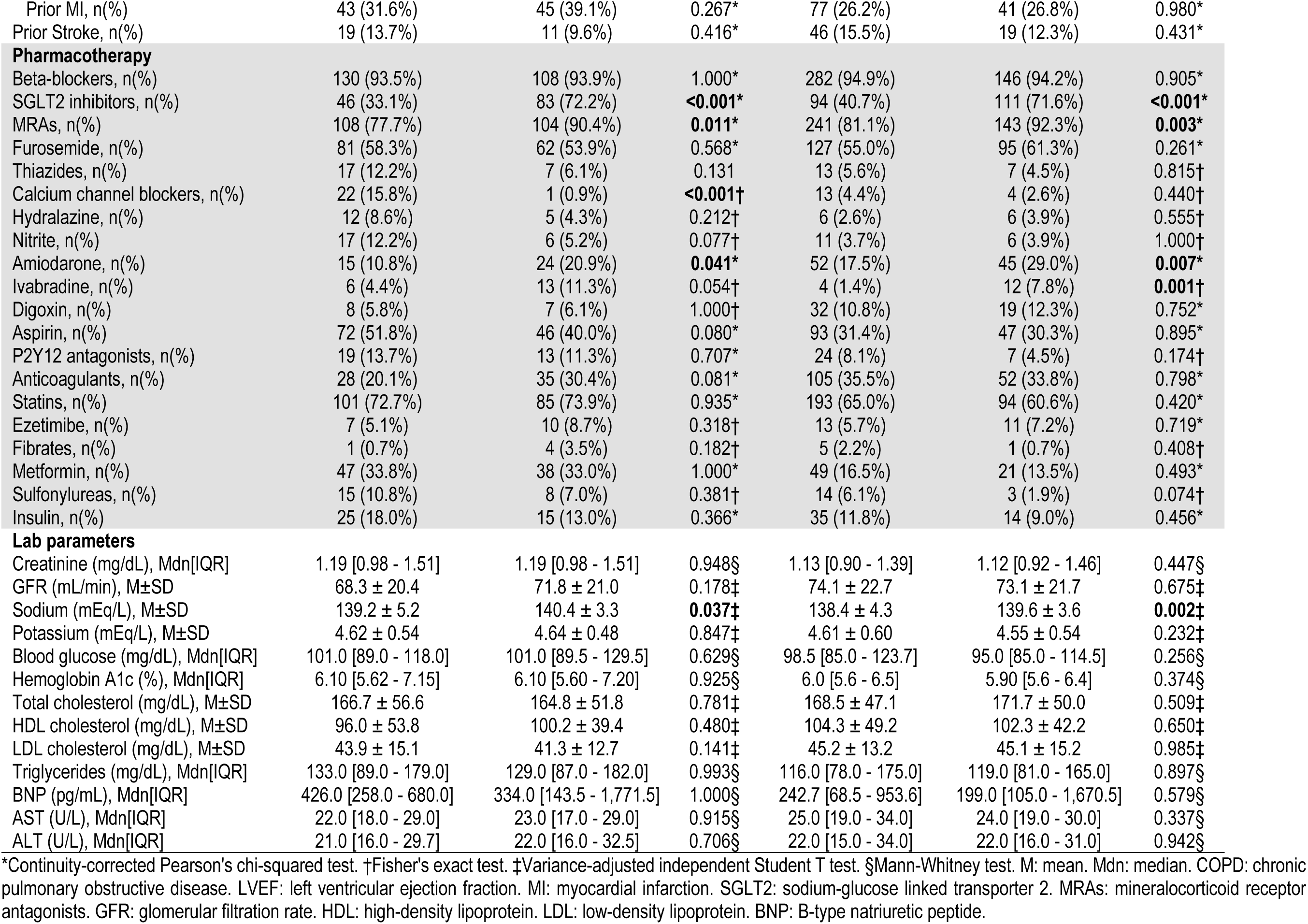
Population characteristics for sacubitril-valsartan compared to either enalapril or losartan.

Regarding HF pharmacotherapy, the sacubitril-valsartan group more often received sodium-glucose cotransporter 2 (SGLT2) inhibitors (72.2 vs. 33.1%, p<0.001), and mineralocorticoid receptor antagonists (MRAs) (90.4 vs 77.7%, p=0.011). These individuals were also less likely to receive calcium channel blockers (CCBs) (0.9 vs 15.8%, p<0.001), and more likely to receive amiodarone (20.9 vs. 10.8%, p=0.041). No significant differences were observed for other drugs (beta-blockers, diuretics, vasodilators, other antiarrhythmics, antidiabetics, lipid-lowering, and antithrombotics). The only statistically significant difference regarding lab parameters was slightly higher natremia in the sacubitril-valsartan group (mean 140.4 vs. 139.2mEq/L), probably related to higher use of MRAs.

### Propensity score matching

Based on clinical knowledge and statistically significant differences in the descriptive analysis, the following covariate candidates were fed to LASSO regression: sex, age, body mass index (BMI), left ventricular ejection fraction (LVEF), glomerular filtration rate (GFR), the classical cardiovascular risk factors (obesity, hypertension, type 2 diabetes mellites, dislipdemia, smoking history), the major HF etiologies (ischemic, hypertensive, Chagas cardiomyopathy, dilated cardiomyopathy, valvular), the other HF drugs (beta-blockers, SGLT2 inhibitors and MRAs), antiarrhythmic drugs, diuretic drugs, antidiabetics, lipid-lowering drugs, antithrombotic drugs, and metabolic lab parameters (hemoglobin A1c, lipid profile). The treatment variable was set as sacubitril-valsartan use, and the outcome variable as combined hospitalization or death.

LASSO cross-validation returned a minimum lambda of 0.052, with following coefficients: LVEF, smoking history, SGLT2 inhibitor use, CCB use, and amiodarone use. Participants were matched based on their propensity score for the use of sacubitril-valsartan. The following covariates were used for the calculation of the propensity score: age, LVEF, smoking history, SGLT2 inhibitor use, MRA use, CCB use, and amiodarone use. The mean standardized difference for distance was 1.27 before matching, and 0.07 after matching (**Supplementary table 1**). The mean standardized difference for covariates ranged from −1.61 to 0.87 before matching, and from −0.07 to 0.07 after matching. Before matching, the population had 115 individuals treated with sacubitril-valsartan 97/103mg twice daily and 139 controls (treated with either enalapril 20mg twice daily or losartan 100mg daily) free from missing data on covariates. The matching process led to a loss of 60 controls, but greatly increased comparability between groups.

### Outcomes

Over the median follow-up of 23.9 months [IQR 11.9 – 41.1], there were 70 primary endpoint events (56 first hospitalizations and 32 deaths). There was a nominal but not statistically significant reduction in combined hospitalization or death in the sacubitril-valsartan group (33.9 vs. 39.7%, p=0.500) (**Table 2**). Similar results were observed for hospitalization (27.8 vs. 30.8%, p=0.779), all-cause mortality (14.8 vs. 19.2%, p=0.536), and 3p-MACE (14.9 vs 16.2%, p=0.972). The notable exception was in NHYA class progression, in which improvement was more frequent in the sacubitril-valsartan group (42.6 vs. 22.1%, p=0.009).

**Table 2.**
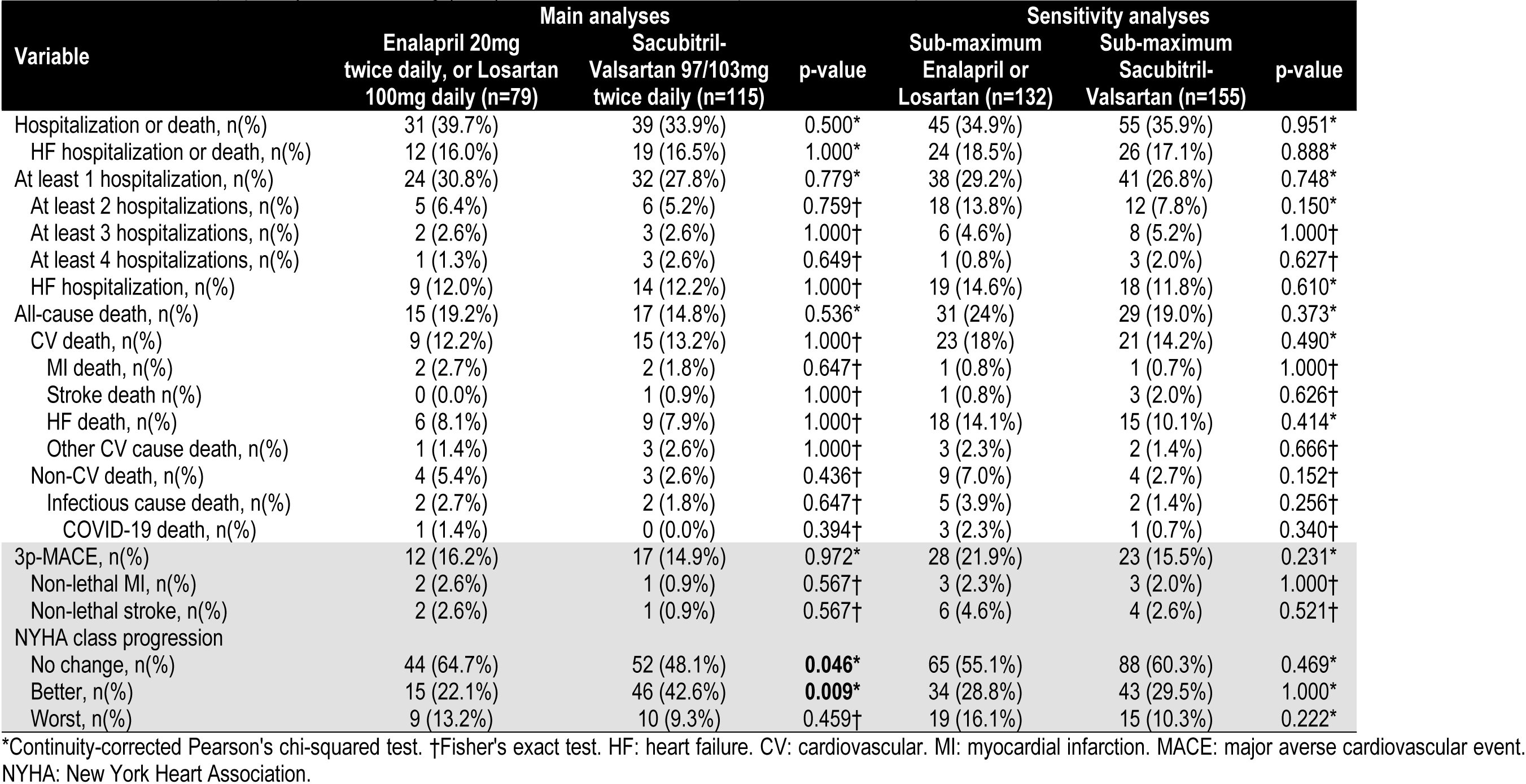
Outcomes after propensity score matching (PSM) for sacubitril-valsartan compared to either enalapril or losartan.

| Variable | Main analyses |  |  | Sensitivity analyses |  |  |
| --- | --- | --- | --- | --- | --- | --- |
|  | Enalapril 20mg<br>twice daily, or Losartan<br>100mg daily (n=79) | Sacubitril-Valsartan 97/103mg<br>twice daily (n=115) | p-value | Sub-maximum<br>Enalapril or<br>Losartan (n=132) | Sub-maximum<br>Sacubitril-Valsartan (n=155) | p-value |
| Hospitalization or death, n(%) | 31 (39.7%) | 39 (33.9%) | 0.500* | 45 (34.9%) | 55 (35.9%) | 0.951* |
| HF hospitalization or death, n(%) | 12 (16.0%) | 19 (16.5%) | 1.000* | 24 (18.5%) | 26 (17.1%) | 0.888* |
| At least 1 hospitalization, n(%) | 24 (30.8%) | 32 (27.8%) | 0.779* | 38 (29.2%) | 41 (26.8%) | 0.748* |
| At least 2 hospitalizations, n(%) | 5 (6.4%) | 6 (5.2%) | 0.759† | 18 (13.8%) | 12 (7.8%) | 0.150* |
| At least 3 hospitalizations, n(%) | 2 (2.6%) | 3 (2.6%) | 1.000† | 6 (4.6%) | 8 (5.2%) | 1.000† |
| At least 4 hospitalizations, n(%) | 1 (1.3%) | 3 (2.6%) | 0.649† | 1 (0.8%) | 3 (2.0%) | 0.627† |
| HF hospitalization, n(%) | 9 (12.0%) | 14 (12.2%) | 1.000† | 19 (14.6%) | 18 (11.8%) | 0.610* |
| All-cause death, n(%) | 15 (19.2%) | 17 (14.8%) | 0.536* | 31 (24%) | 29 (19.0%) | 0.373* |
| CV death, n(%) | 9 (12.2%) | 15 (13.2%) | 1.000† | 23 (18%) | 21 (14.2%) | 0.490* |
| MI death, n(%) | 2 (2.7%) | 2 (1.8%) | 0.647† | 1 (0.8%) | 1 (0.7%) | 1.000† |
| Stroke death n(%) | 0 (0.0%) | 1 (0.9%) | 1.000† | 1 (0.8%) | 3 (2.0%) | 0.626† |
| HF death, n(%) | 6 (8.1%) | 9 (7.9%) | 1.000† | 18 (14.1%) | 15 (10.1%) | 0.414* |
| Other CV cause death, n(%) | 1 (1.4%) | 3 (2.6%) | 1.000† | 3 (2.3%) | 2 (1.4%) | 0.666† |
| Non-CV death, n(%) | 4 (5.4%) | 3 (2.6%) | 0.436† | 9 (7.0%) | 4 (2.7%) | 0.152† |
| Infectious cause death, n(%) | 2 (2.7%) | 2 (1.8%) | 0.647† | 5 (3.9%) | 2 (1.4%) | 0.256† |
| COVID-19 death, n(%) | 1 (1.4%) | 0 (0.0%) | 0.394† | 3 (2.3%) | 1 (0.7%) | 0.340† |
| 3p-MACE, n(%) | 12 (16.2%) | 17 (14.9%) | 0.972* | 28 (21.9%) | 23 (15.5%) | 0.231* |
| Non-lethal MI, n(%) | 2 (2.6%) | 1 (0.9%) | 0.567† | 3 (2.3%) | 3 (2.0%) | 1.000† |
| Non-lethal stroke, n(%) | 2 (2.6%) | 1 (0.9%) | 0.567† | 6 (4.6%) | 4 (2.6%) | 0.521† |
| NYHA class progression |  |  |  |  |  |  |
| No change, n(%) | 44 (64.7%) | 52 (48.1%) | **0.046\*** | 65 (55.1%) | 88 (60.3%) | 0.469* |
| Better, n(%) | 15 (22.1%) | 46 (42.6%) | **0.009\*** | 34 (28.8%) | 43 (29.5%) | 1.000* |
| Worst, n(%) | 9 (13.2%) | 10 (9.3%) | 0.459† | 19 (16.1%) | 15 (10.3%) | 0.222* |

\*Continuity-corrected Pearson's chi-squared test. †Fisher's exact test. HF: heart failure. CV: cardiovascular. MI: myocardial infarction. MACE: major adverse cardiovascular event. NYHA: New York Heart Association.

ATET analysis confirmed these findings, showing no significant reductions for the primary outcome (OR 0.650, 95%CI 0.354 – 1.195, p=0.165), or its components. It also revealed an even stronger association for NYHA class improvement (OR 3.902, 95%CI 1.745 – 8.726, p=0.001) (**Table 3**).

**Table 3.** Average treatment effect on the treated (ATET) for sacubitril-valsartan compared to either enalapril or losartan (maximum dosages).

| Variable | Main analyses |  |  |  | Sensitivity analyses |  |  |  |
| --- | --- | --- | --- | --- | --- | --- | --- | --- |
| | $\beta$ | SE | p-value | OR (95%CI) | $\beta$ | SE | p-value | OR (95%CI) |
| Hospitalization or death | -0.431 | 0.311 | 0.165* | 0.650 (0.354 - 1.195) | 0.202 | 0.271 | 0.455* | 1.224 (0.702 - 2.081) |
| HF hospitalization or death | -0.236 | 0.391 | 0.546* | 0.790 (0.367 - 1.700) | 0.025 | 0.336 | 0.941* | 1.025 (0.531 - 1.980) |
| At least 1 hospitalization | -0.286 | 0.324 | 0.377* | 0.751 (0.398 - 1.418) | 0.001 | 0.290 | 0.998* | 1.001 (0.567 - 1.765) |
| At least 2 hospitalizations | -0.254 | 0.628 | 0.686* | 0.776 (0.227 - 2.657) | -0.667 | 0.399 | 0.094* | 0.513 (0.235 - 1.121) |
| HF hospitalization | -0.428 | 0.421 | 0.310* | 0.652 (0.286 - 1.488) | -0.230 | 0.367 | 0.531* | 0.795 (0.387 - 1.631) |
| All-cause death | -0.214 | 0.410 | 0.602* | 0.808 (0.362 - 1.802) | -0.049 | 0.321 | 0.879* | 0.952 (0.508 - 1.786) |
| CV death | 0.057 | 0.465 | 0.902* | 1.059 (0.426 - 2.635) | -0.127 | 0.351 | 0.716* | 0.880 (0.443 - 1.751) |
| HF death | 0.523 | 0.660 | 0.428* | 1.688 (0.463 - 6.154) | -0.226 | 0.396 | 0.568* | 0.797 (0.367 - 1.733) |
| 3p-MACE | -0.193 | 0.415 | 0.642* | 0.824 (0.365 - 1.861) | -0.268 | 0.328 | 0.415* | 0.765 (0.402 - 1.456) |
| NYHA class progression |  |  |  |  |  |  |  |  |
| No change | -0.942 | 0.338 | **0.005\*** | 0.390 (0.201 - 0.757) | 0.153 | 0.266 | 0.564* | 1.165 (0.692 - 1.962) |
| Better | 1.362 | 0.411 | **0.001\*** | 3.902 (1.745 - 8.726) | 0.212 | 0.300 | 0.480* | 1.236 (0.687 - 2.223) |
| Worst | -0.427 | 0.491 | 0.385* | 0.653 (0.249 - 1.710) | -0.660 | 0.367 | 0.072* | 0.517 (0.252 - 1.061) |

\*Weighted logistic regression and log odds (beta). HF: heart failure. CV: cardiovascular. 3p-MACE: 3-point major adverse cardiovascular event. NYHA: New York Heart Association.

### Sensitivity analysis

We also examined data from the 452 patients receiving any non-maximal dosages of the investigated drugs. This population was 61.5% male, with a mean age of 61.4 years, and a median follow-up of 19.3 [IQR 8.7 – 33.7] months. A total of 155 patients (34.2%) used sacubitril-valsartan, 201 (44.4%) enalapril, and 96 (21.2%) losartan. Differences in population characteristics stratified by drug were similar to those seen with optimal dosages, with a lower EF and more frequent use of SGLT2 inhibitors, MRAs and amiodarone in the sacubitril-valsartan group. After PSM (**Supplementary table 2**), no differences were observed for the primary or secondary outcomes, which was confirmed by ATET.

## DISCUSSION

This study used real-world data from the BEAT-HF cohort to compare the clinical outcomes of patients with HFrEF prescribed the guideline-recommended maximum dosages of sacubitril-valsartan (97/103mg twice daily) versus either enalapril (20mg twice daily) or losartan (100mg daily). No significant reductions were observed in the primary outcome, combined hospitalization or death (OR 0.650, 95%CI 0.354 – 1.195, p=0.165), nor in its components, first hospitalization (OR 0.751, 95%CI 0.398 – 1.418, p=0.377) and all-cause mortality (OR 0.808, 95%CI 0.362 – 1.802, p=0.602). Our findings weren’t able to support reductions in hospitalization or mortality for sacubitril-valsartan compared to enalapril or losartan at guideline-recommended maximum dosages.

Following the PARADIGM-HF trial, subsequent studies comparing sacubitril-valsartan with equivalent dosages of ACE inhibitors or ARBs found no statistically significant differences between groups in patients with either preserved or reduced EF [11–13]. In the LIFE trial, among patients with advanced HFrEF, sacubitril-valsartan failed to further reduce NT-proBNP compared to Valsartan (AUC 0.95, 95%CI 0.84 – 1.08, p=0.45) [11]. Similarly, the PARADISE-MI trial, which enrolled patients with left ventricular dysfunction following myocardial infarction, did not demonstrate superiority of sacubitril-valsartan over Ramipril in preventing cardiovascular death or HF events (HR 0.90, 95%CI 0.78 – 1.04, p=0.17) [12]. Furthermore, a recent meta-analysis compared sacubitril-valsartan with ACE inhibitors and ARBs at equivalent dosages and found no significant reductions in all-cause mortality (RR 0.90, 95%CI 0.76 – 1.07, p=0.38, i²=0%) [13]. These studies also support the hypothesis that the favorable outcomes of sacubitril-valsartan in the PARADIGM-HF trial may have been influenced, at least partially, by the use of a suboptimal control regimen.

Despite the absence of a significant reduction in hospitalization or mortality, we observed a strong association of sacubitril-valsartan with improved NYHA class (OR 3.902, 95%CI 1.745 – 8.726, p=0.001), probably attributable to its intrinsic diuretic effect. It is noteworthy that no significant differences were observed in diuretic use between the groups, and that PSM achieved near-perfect balance for SGLT2 inhibitors (72.1% vs. 70.0%) and MRAs (90.4% vs. 91.1%). This hypothesis was further tested through multivariate logistic regression (accounting for age, sex, FE, GFR, HF drugs, and diuretics), which showed similar results (sacubitril-valsartan w/ an OR of 2.805, 95%CI 1.275 – 6.172, p=0.010) (**Supplementary table 3**). This is consistent with other studies showing improvement in NYHA class and quality of life scores, despite no significant reductions in hospitalization or mortality [4,13].

A final topic to be addressed is the pharmacoeconomics of sacubitril-valsartan. Considering the guideline-recommend maximum dosages and retail prices found online by the investigators in United States pharmacies, sacubitril-valsartan costs approximately 600 USD per month, whereas Enalapril and Losartan are a lot more affordable, at around 25 and 10 USD per month, respectively. In Brazil, despite the generic drug policy – which allows low-cost production of medications – sacubitril-valsartan remains considerably more expensive, costing approximately 55 USD per month, versus Enalapril and Losartan, available for as little as 2 and 3 USD per month, respectively. No formal pharmacoeconomic analyses were performed in this study; However, given the lack of significant reductions in hard outcomes compared to enalapril or losartan in our population, our findings raise concerns about the cost-effectiveness of sacubitril-valsartan in resource-limited settings.

The findings of this study underscore the importance of incorporating real-world evidence into the evaluation of therapeutic strategies. In an increasingly complex landscape marked by multiple pharmacological options, understanding how new therapies perform outside the controlled environment of clinical trials is essential. Sacubitril-valsartan should be assessed not only for its efficacy, but also in terms of applicability and cost-effectiveness – particularly in low-and middle-income countries (LMICs) with constrained healthcare resources, such as Brazil.

### Limitations

The main limitation of our study is its relatively small sample size. The nominal reduction in the primary outcome observed in the sacubitril-valsartan group raises the question of whether this difference would reach statistical significance in a larger cohort. Using GPower 3.1.9.7, the statistical power (1 - β error) for the measured outcomes was excellent (0.893). This supports the argument that our sample size is sufficient to measure outcomes, and that the reduction in hospitalization or death with sacubitril-valsartan is in fact small compared to enalapril or losartan at guideline-recommended maximum dosages.

The observational and partially retrospective design is another limitation. To account for baseline differences between treatment groups, we performed propensity score matching (PSM). Without this adjustment, the higher prevalence of SGLT2 inhibitors (72.2 vs. 33.1%, p<0.001) and MRAs (90.4 vs. 77.7%, p=0.011) in the sacubitril-valsartan group could have substantially influenced outcomes.

The long follow-up period was overall positive as it allowed for more end-point events to happen. However, the start of the observation period in August 2010 creates a potential bias, as the sacubitril-valsartan group would be more recent, having better access to modern therapies for HF (ie, SGLT2 inhibitors) and other diseases. This problem was partially addressed by PSM. To further isolate the impact of sacubitril-valsartan, we also estimated the average treatment effect on the treated (ATET).

## CONCLUSION

In this real-world study of patients with heart failure with reduced ejection fraction, sacubitril-valsartan did not demonstrate a significant reduction in the combined outcome of hospitalization or death when compared to guideline-recommended maximum dosages of either enalapril or losartan. Likewise, no significant benefits were observed for the individual outcomes of hospitalization, all-cause mortality, or major adverse cardiovascular events (3p-MACE).

Despite the lack of impact on these key clinical events, treatment with sacubitril-valsartan was strongly associated with an improvement in patients’ functional status, as assessed by the NYHA classification.

These findings suggest that the clinical benefits of sacubitril-valsartan on mortality and hospitalization may be more modest than reported in previous pivotal trials. The results highlight the need for further research to clarify the role of sacubitril-valsartan in specific patient subgroups and to evaluate its cost-effectiveness in various real-world settings.

## DECLARATIONS

### Ethics approval and consent to participate

This study was conducted in accordance with the Declaration of Helsinki and approved by the Institutional Review Boards of Instituto de Gestão Estratégica de Saúde do Distrito Federal (IGESDF; approval numbers 35543020.1.1001.8153 and 35543020.1.0000.8153) and Fundação de Ensino e Pesquisa em Ciências da Saúde (FEPECS; approval number 72838823.5.0000.5553). Informed consent was obtained from prospectively enrolled participants. For retrospectively enrolled patients, including those deceased or unreachable, a waiver of consent was granted by the IRBs. Copies of the informed consent forms (both written and online versions) have been provided as supplementary files.

### Consent for publication

Not applicable.

### Clinical trial number

Not applicable.

### Funding

This research received no external funding.

### Competing interests

The authors declare that they have no known competing financial interests or personal relationships that could have appeared to influence the work reported in this paper.

### Availability of data and materials

The datasets generated and/or analyzed during the current study are available from the corresponding author on reasonable request.

### Authors’ contributions

**Conceptualization:** Renato C. Barros; Luiz Sérgio F. de Carvalho.

**Gathering data:** Vinícius C. Fiusa; Gabrielle N. Tim; Victoria T. V. Goulart; Caio Henrique M. Nascimento; Gustavo T. de Moura; Adriana J. B. A. Guimarães; Renato C. Barros.

**Statistical analysis:** Vinícius C. Fiusa.

**Writing of the manuscript:** Renato C. Barros; Vinícius C. Fiusa.

**Review of scientifically-important content:** Luiz Sérgio F. de Carvalho; Filipe A. Moura; Sergio Henrique Rodolpho Ramalho; Armindo Jreige Jr; Adriana J. B. A. Guimarães.

**Supervision:** Luiz Sérgio F. de Carvalho.

All authors read and approved the final manuscript.

## Data Availability

All relevant data are within the manuscript and its Supporting Information files.

## Notes

### Competing Interest Statement

The authors have declared no competing interest.

### Funding Statement

The author(s) received no specific funding for this work.

### Author Declarations

This study was conducted in accordance with the Declaration of Helsinki and approved by the Institutional Review Boards of Instituto de Gestão Estratégica de Saúde do Distrito Federal (IGESDF approval numbers 35543020.1.1001.8153 and 35543020.1.0000.8153) and Fundação de Ensino e Pesquisa em Ciências da Saúde (FEPECS approval number 72838823.5.0000.5553). Informed consent was obtained from prospectively enrolled participants. For retrospectively enrolled patients, including those deceased or unreachable, a waiver of consent was granted by the IRBs. Copies of the informed consent forms (both written and online versions) have been provided as supplementary files.

